# Comparison of cough particle exposure for indoor commercial and aircraft cabin spaces

**DOI:** 10.1101/2021.03.24.21254275

**Authors:** Angela C. Davis, Danny J. Menard, Andrew D. Clark, Joshua J. Cummins, Nels A. Olson

## Abstract

To compare the transport of respiratory pathogens, computational fluid dynamics (CFD) simulations were performed to track particles released by coughing from a passenger on a Boeing 737 aircraft, and by a person in a comparable indoor commercial space. Simulation data were post-processed to calculate the amounts of particles inhaled by nearby persons in both environments. The effects of different airflow rates, placement of air inlets, positioning and distances between index (coughing) and susceptible (inhaling) persons were also analyzed. The removal of airborne particles from the indoor environment, due ventilation and deposition onto surfaces, was compared to that of an aircraft cabin. In an aircraft cabin 80% of the particles were removed 5 to 12 times faster than in the indoor commercial space; ultimately resulting in 7 times less particulate mass inhaled in the aircraft cabin.

## Introduction

Of the industries affected by the SARS-CoV-2 (COVID-19) pandemic, few were more impacted than the air travel industry that experienced a 95% drop in passengers compared to 2019 [1]. As segments of the population continued to travel, questions began to emerge regarding the relative risk of disease transmission during air travel and, specifically, within the aircraft cabin due to the close-quarters seating.

A computational fluid dynamic study of cough particulate dispersal and inhalation in an aircraft cabin was performed by Davis et al. [2]. The study found an average exposure of 0.05% of the particulate mass emitted by a cough with a maximum exposure of 0.3%. A number of researchers have performed CFD studies of cough exposure in indoor commercial spaces (ICS); however, the cough dynamics, model inputs, and methods used differed from each other, and from those previously reported by Davis et al. for an aircraft [3, 4, 5]. To understand the exposure and relative risk of exposure in an aircraft cabin, a direct comparison CFD analysis was performed for an ICS using consistent methods and inputs with the Davis et al. aircraft cabin study.

A multi-layered approach is required to control the aerosol and small droplet particles responsible for the spread of respiratory disease. The multi-layered approach has proven valuable in many environments. Physical distancing, ventilation, face coverings, personal protective equipment, frequent hand washing and surface disinfection are all important additional components in the control of a novel respiratory disease. As with any redundancy factor for the improvement of reliability, no one control on its own can be as effective as all controls combined.

A well-designed Environmental Control Systems (ECS) is one of those layers. An ECS controls the number of air exchanges per hour (ACH) and creates airflow patterns that direct particles away from occupants. A typical ICS has 2 to 5 air changes per hour, which is considerably fewer than a typical aircraft cabin [6]. Additionally in an ICS, the airflow pattern tends to be less precisely defined than in an aircraft cabin, because of the variability in room geometry and occupant position relative to air inlets and outlets.

The ECS on a typical passenger aircraft generates approximately twenty to thirty ACH when calculated for an empty cabin [7]. As the volume of the cabin is decreased by filling it with passengers and luggage, the ACH rate notably increases. The cabin air typically consists of 50% outside air and 50% recirculated air. The recirculated air is passed through high-efficiency particulate air (HEPA) filters to remove particles, including those carrying pathogens, at an efficacy of 99.97% at the most penetrating particle size of 0.3 μm (i.e., the particle size that most easily escapes the filter unit). Additionally, by directing air flow to move from top to bottom of the cabin, rather than from front to back, further limits particle spread within the cabin (Figure 1). The particulate exposure of neighboring passengers is reduced by positioning of passengers facing forward (rather than facing each other) for the majority of the flight time. The high back seat design also acts as a barrier, similar to Plexiglas™ barriers commonly seen in ICS, and helps prevent direct transmission of droplets.

**Figure 1.**
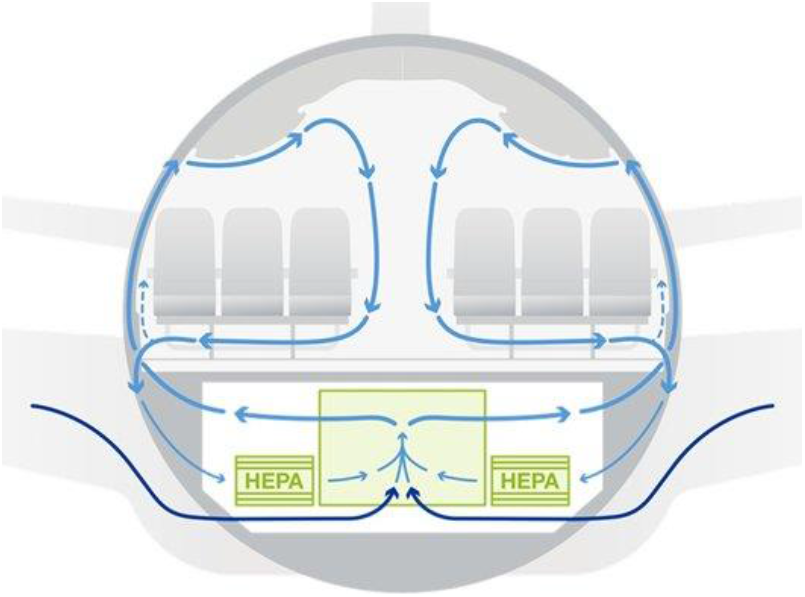
Design for airflow in the passenger cabin.

To characterize the differences between tightly controlled environments like aircraft and more general indoor environments, CFD simulations were performed that tracked particles released by a single cough in an ICS. Common seating arrangements, air inlet and outlet configurations, and room geometries were used for the CFD models. The number, size and rate of release for cough particles was held constant between the ICS and airplane models. To allow for direct comparisons, the methods and results for an aircraft cabin presented herein are replicated from Davis (2021) [2]. Ratios between the two models were calculated, and comparisons made between the times required for particle removal. Further comparisons were made for the mass inhaled by nearby persons in each environment. Finally, the overall distribution of mass inhaled within each environment, and the effect of distance from the index cougher was compared.

## Methods

### CFD Overview

Simulations and analyses were performed using the method described by Davis (2021) [2]. CFD simulations were performed using Ansys Fluent™ 19.2 computational fluid dynamics simulation software for the aircraft (737 Boeing Sky Interior cabin) and Ansys Fluent™ 2020R2 for the indoor commercial space. Airflow and thermal boundary conditions are listed in Table 1, with the values of ACH calculated based on an unoccupied and unfurnished environment.

**Table 1.** Airflow and thermal boundary conditions used for CFD simulations.

|  | **Aircraft** | **Indoor Commercial Space** |
| --- | --- | --- |
| Supply flow rate [Actual m<sup>3</sup>/min] | 9.15 to 16.65 | 4.84 to 19.34 |
| Return flow rate [Actual m<sup>3</sup>/min] | Same as supply | Same as supply |
| Air changes per hour | 24.7 to 44.9 | 2 to 8 |
| Relative humidity [%] | 0 to 20 | 50 |
| Occupant heat generation [W/person] | 70 | 70 |
| Walls [°F] | 55 to 65 | Adiabatic |
| Ceiling, floor, stowage bins | Adiabatic | Adiabatic |
| Front and back interfaces | Periodic | Non-periodic |
| Supply air temperature [°F] | 62 to 67 | 65 |
| Environment average temperature [°F] | 75 to 77 | 70 to 80 |

In each simulation, a single cough was emitted by an index cougher over the course of seconds, using a time-dependent flow rate [8] and a particle size distribution including sizes down to 0.1 μm [9]. Expiratory particles were simulated using the density of water, a volatile fraction of 90% [10], and evaporated to droplet nuclei over approximately 2 seconds using a vapor pressure corrected for the effect of lung surfactants, mucin protein and sodium chloride [11]. The rate of evaporation in the aircraft vs. the ICS was different due to differences in the relative humidity, however, the time to reach the droplet nuclei size was comparable and on the order of a few seconds between both methods. When particles impacted surfaces, they were assumed captured and were removed from the airborne part of the simulation as deposited material.

Manikins (not wearing masks) were used in the simulations to provide physical obstacles and generate heat, which should result in more realistic airflow patterns than a measurement volume without manikins. For each manikin used in the simulation, a breathing zone was defined as a cube with side length of 0.305 m. (Supplementary Figure S1) and a volume of 0.0227 m^3^, after subtracting the volume representing the subject’s body. For each manikin, the mass of particles in their breathing zone was tracked throughout the simulations, and the inhaled mass was calculated by applying the inhalation portion of a tidal breathing function as previously described by Gupta et al. [12].

The mass inhaled in each environment was calculated over the duration of the simulation. ICS simulations were simulated for 15 minutes regardless of air flow rate. At 4 ACH in the ICS, approximately 19% of the particles remained airborne at the end of the simulated period. The aircraft CFD models were allowed to run until 1% or less of the particles remained in the air, which ranged from 4 to 7.5 minutes of simulation time depending on air flow rate. Overall, mass remaining airborne in the ICS simulations should lead to a slight underestimation of the inhaled mass when compared to an aircraft. The underestimation in mass inhaled in an ICS is expected to be small because the particles that remained airborne after 15 minutes have a small mass and volume, and can only carry a small amount of nonvolatile material; all resulting in a comparatively small addition to the inhaled mass in the ICS.

The amount of material inhaled was quantified in terms of nonvolatile mass, defined as the mass of particles after evaporation, and expressed as a percentage of the nonvolatile mass expelled by the index subject. The total nonvolatile mass, inhaled in the ICS over the duration of the CFD simulations was calculated for each seat except the infectious index cougher. The nonvolatile mass inhaled was not calculated for the index cougher because the particles were not fully evaporated while in the index cougher’s breathing zone.

### Aircraft Simulations

A five-row section of the 737 Boeing Sky Interior (BSI) cabin was used for aircraft simulations, with the seat chart shown in Figure 2(a).

**Figure 2.**
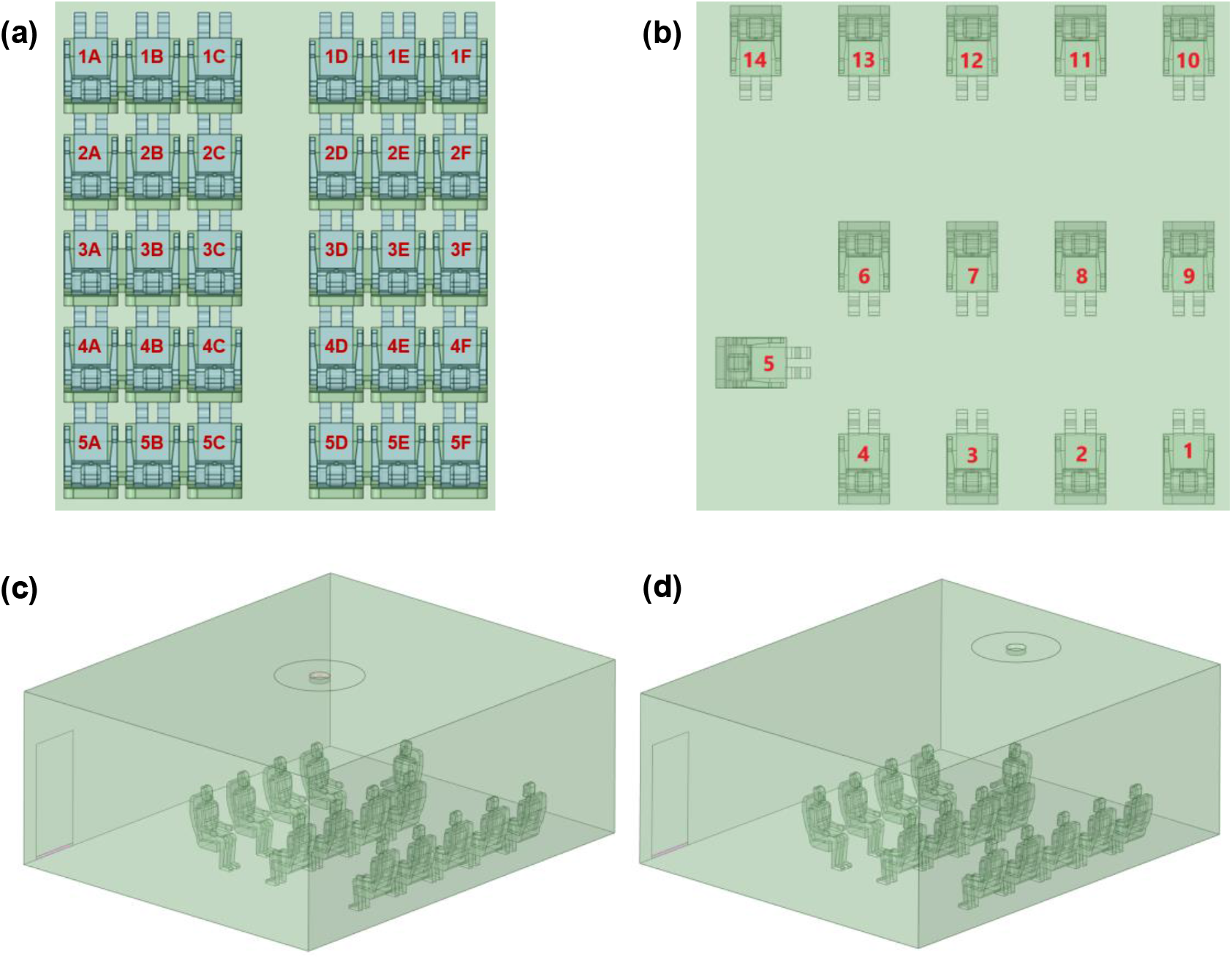
Simulation geometries. **(a)** Airplane seat positions, **(b)** ICS seat positions, **(c)** ICS with a center inlet position, **(d)** ICS with an offset inlet position.

The cases analyzed previously by Davis et al. [2] and listed in Supplementary Table S1 were used for comparison to the ICS model. To account for the effects of random low frequency fluctuations in airflow as described earlier by Davis’s team, several time points from the steady state solutions were used as the initial conditions (Supplementary Table S1) for one of the index cougher positions upon a cough expiratory event. For the purposes of this study, the Personal Air Outlets (PAOs) common on most commercial aircraft were all in the closed position as analysis of the PAO effects did not yield a clear recommendation for their use.

### Indoor Commercial Space Simulations

The indoor commercial space geometry selected was not intended to represent any one specific type of building or scenario, but rather to meet the minimum airflow rates for a range of indoor commercial spaces as defined in ASHRAE 62.1-2019. Minimum requirements for airflow rates in ICS, vary from 0.06 to 0.18 cubic feet per minute (cfm)/ft^2^ area and 5 to 10 cfm/occupant depending on the and building and room. The airflow rates in this study were selected to cover a wide range of potential environments, and varied from 0.3 to 1.3 cfm/ft^2^ (with an occupant density of 25 to 150 people/1000 ft^2^).

Due to the large number of particles released from a single cough, the CFD simulations performed were computationally expensive, which resulted in a need to limit the variables evaluated. The ICS simulation was limited to a single air inlet/outlet pair; had a size that would allow several positions for the index cougher and susceptible inhaler, and allowed the orientation of the occupants to be described in easily understood ways e.g., facing one another, downwind, or in the plume of the cough. The number of manikins positioned within the space enabled tracking of particles within breathing zones throughout the environment which captured variability while limiting the number of required simulations.

Fourteen manikins were arranged around the ICS in rows (Figure 2(b)) and in a seated position to provide a comparison to an aircraft cabin. The dimensions of the ICS were 7.62m x 6.25 m x 3.05 m. An air outlet with dimensions of 5.08 cm x 91.44 cm was placed in the corner of the room behind seat 1 (Figure 2(c,d)), to represent a return air grille. To accurately model the relatively small but geometrically complex ceiling air-supply-diffuser in the room and to accelerate the simulation, a validated simplified method by Mohammed (2013) [13] was adopted for this study.

For the ICS, the supply air was assumed to be free of contaminates, and the study did not consider the recirculation of air or expiratory particles. Recirculation of cough particles may increase occupant exposure, but the added complexity of including recirculation in the ICS simulation would likely have had a small effect compared to exposure to the concentrated cough plume. The exclusion of recirculation from the ICS adds to the conservative approach for the aircraft comparison.

Nine simulations were performed for the ICS (Supplementary Table S2). To understand the effect of the index cougher’s location within the indoor space, three positions were tested, with index coughers located in seats 4, 5 and 12 (Figure 2(b)). One simulation was performed with seats 6-9 removed from the geometry to observe unobstructed particle transport for the index cougher located in seat 4. Different air inlet positions were studied to observe variability based on the design of the ICS. For each cough position, two air inlet positions were analyzed, one with a diffuser air inlet positioned above the center of the room (Figure 2(c)), and the other with a diffuser air inlet positioned off-center directly above seat 5 (Figure 2(d)). The effect of supply air flow rate was also studied for an index cougher located in seat 4 with a center inlet position.

The initial air flow pattern for three conditions in the ICS simulation are shown in Figure 3. The Figure shows the effect of varying the air flow rate, a similar figure appears in the Davis et al. 2021 CFD analysis of an aircraft [2]. The 4 ACH condition was used to generate the majority of the data because it is an industry standard and a common ACH for ICS environments. The 2 ACH and 8 ACH conditions were chosen as a comparison to 4 ACH and to show the difference in the time required to remove cough particles from the overall ICS. The 2 ACH flow condition (Figure 3(a)) simulated a low air flow rate, however, the cool supply air fell to the floor and did not efficiently mix with the room air. The air flow conditions at 4 and 8 ACH (Figure 3(b, c)) appeared to be well-mixed, with the highest air flow velocities along the ceiling and walls. The direction of the initial air flow impacted the initial direction of the cough plume. At 8 ACH, the initial air flow field at seat 4 was directed towards the ceiling and at 4 ACH with a center inlet, the initial air flow at seat 4 was directed towards seats 7 to 9. Different initial air flow conditions at each air flow rate were not studied for the ICS, but are likely to have an effect on the mass inhaled for a given occupant by changing the initial direction of the cough plume.

**Figure 3.**
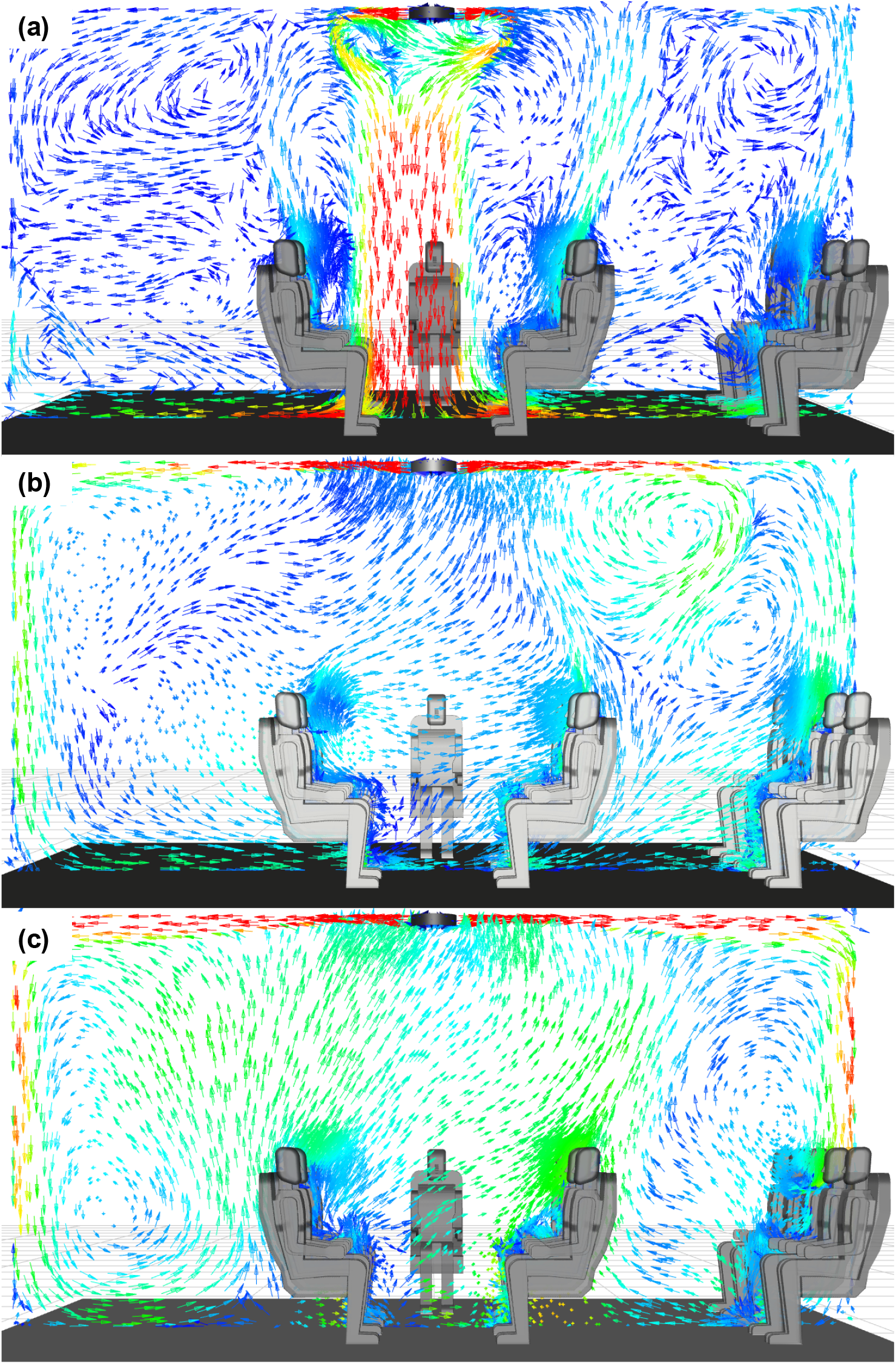
Velocity vectors of an indoor commercial space at **(a)** 2 ACH, **(b)** 4 ACH and **(c)** 8 ACH with a center inlet. Red arrows indicate highest velocity air, yellow to green arrows indicate intermediate air velocities, and teal to dark blue arrows indicate lowest air velocities. All are presented on a linear scale.

## Results and Discussion

### Particle Removal Dynamics

Particles were removed from the airborne environment through air outlets and by deposition onto surfaces. In both of the CFD environments, the decay in the number of expiratory particles over the course of the simulation was monitored (Figure 4). The initial features on the decay curves correspond to particle deposition on surfaces. After the release of particles, the majority of deposition occurred within the first 30 seconds in an aircraft cabin, and within the first two minutes in the ICS.

**Figure 4.**
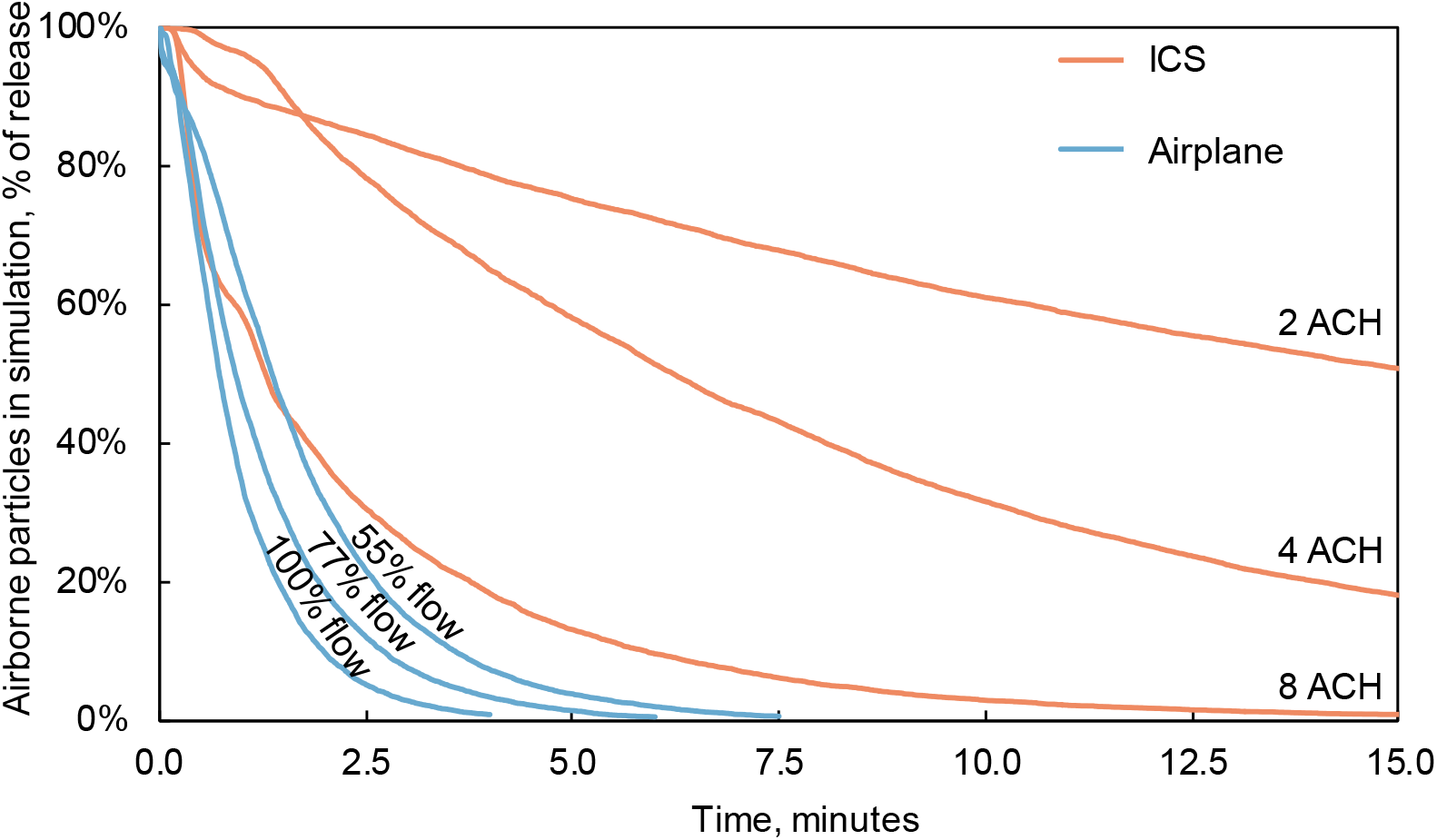
Decay of total expiratory particles vs. time after initial release at 2, 4 and 8 ACH in the ICS compared to 100%, 77%, and 55% supply air flow in aircraft cabin.

Particles were removed more quickly from the aircraft cabin than the ICS. In the ICS at the conditions for a constant 4 ACH, the position of the air supply inlet, and index cougher affected the particle removal dynamics. Within approximately 12.4 to 15 minutes, 80% of the particles were removed from the air at 4 ACH. For an aircraft cabin at 100% flow rate, the index location, initial air flow condition, and relative humidity did not considerably impact particle removal, with 80% of the particles removed within 1.2 to 1.4 minutes.

In both the ICS and aircraft environments, the supply airflow rate had a large effect on particle removal (Figure 4). At higher airflow rates, particles were more rapidly removed. For the ICS, increasing the supply airflow rate to 8 ACH reduced the time for 80% particle removal 3.7 minutes (95% in 7.8 min, 99% in 12.5 min). When the air supply flow rate was decreased to 2 ACH, the time for particle removal increased, with a 50% removal at approximately 15 minutes, and 80% removal beyond the 15 minute simulation time. For an aircraft cabin with an index in seat 4, depending on supply airflow rate, 80% of particles were removed in 1.3 to 2.6 minutes (95% in 2.3 to 4.5 min, 99% in 3.3 to 6.3 min). When compared to the ICS, 80% of particles were removed 5 to 12 times more rapidly in the aircraft cabin, depending on the air flow rate.

### Inhaled Mass

The most important consideration for preventing the transmission of disease is the mass of particles inhaled by a susceptible person after an index person’s cough. It is assumed that the total nonvolatile mass inhaled corresponds to the relative risk of infection. However, different particle sizes may carry different viral loads, which can impact the risk of infection [14, 15]. As it is currently unknown which particle sizes are the highest risk for COVID-19, all particle sizes inhaled were grouped together to calculate the total mass inhaled for each occupant.

The comparisons of inhaled mass between environments are presented for 4 ACH in the ICS, and 100% air flow in the airplane. Comparisons were not made for other flow rates because factors impacting variability in inhaled mass within each environment, such as index position, were only studied at 4 ACH in the ICS and 100% air flow in the airplane. All of the simulations performed at a common supply air flow rate are combined in the following comparisons, unless otherwise stated. This method of comparison skews the average and median results in an aircraft cabin to that of an index in seat 3D; however, the minimum and maximum values observed remain representative for all index positions.

In the airplane, more occupants inhaled less mass than in the ICS and the relative frequency of nonvolatile mass inhaled is shown in Figure 5 with bin widths of 0.025% of the total nonvolatile mass released. The relative frequency is the number of occupants who inhaled mass within each bin, divided by the total number of occupants studied for each environment, N. In the airplane, 93% of the occupants were exposed to less than 0.1% of the mass released, whereas 95% of the ICS occupants were exposed to more than 0.1%. Overall, when comparing the minimum and maximum, mean, and median mass inhaled, as shown in Table 2, exposure in the airplane was 7 to 8 times lower than in the indoor commercial space.

**Table 2.** Comparison of nonvolatile mass inhaled (% of nonvolatile mass released) in an aircraft cabin at 100% supply air flow rate to an indoor commercial space at 4 ACH, presented as a ratio (right hand column)

| **Nonvolatile Mass Inhaled** | **Aircraft Cabin 100% Flow Rate** | **Indoor Commercial Space 4 ACH** | **Nonvolatile Mass Inhaled Ratio: ICS/Aircraft** |
| --- | --- | --- | --- |
| Minimum | 0.009% | 0.07% | 8 |
| Maximum | 0.3% | 2% | 7 |
| Mean | 0.04% | 0.3% | 7 |
| Median | 0.03% | 0.2% | 8 |

**Figure 5.**
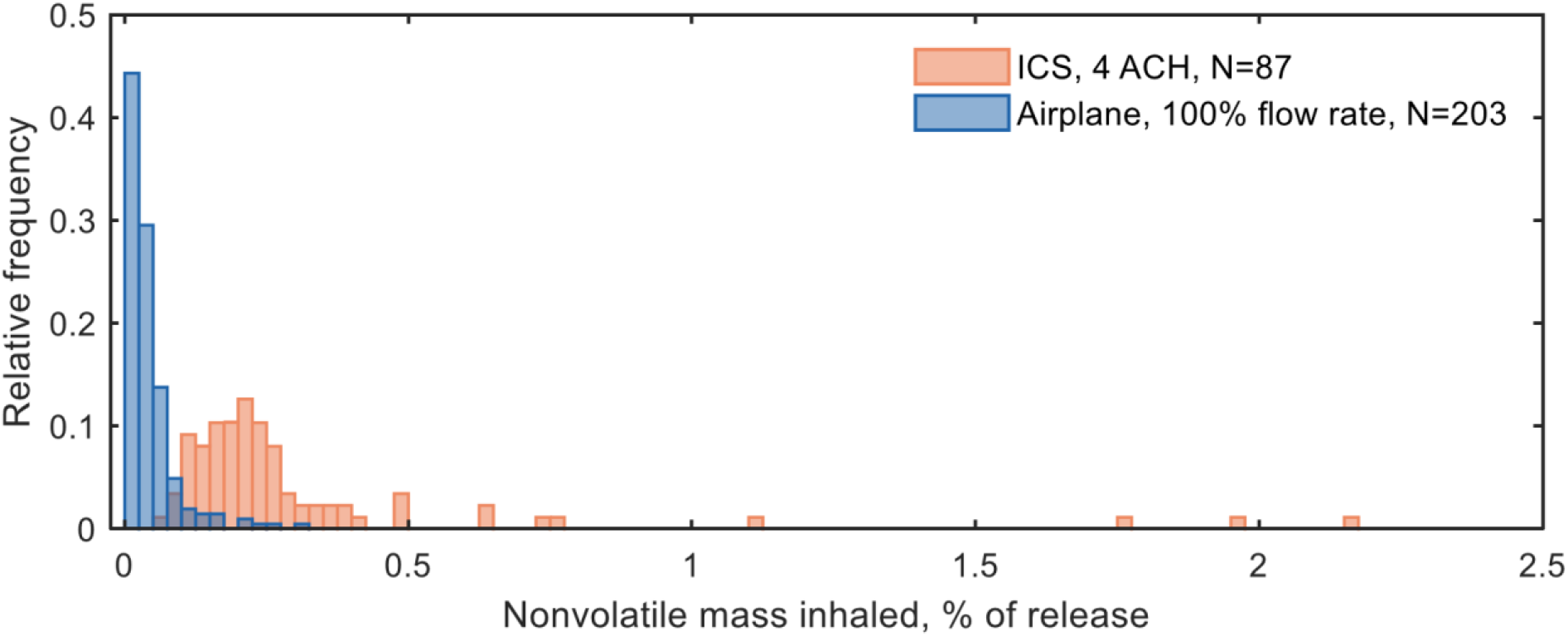
More occupants in the airplane were exposed to less mass than in the ICS. Distribution of exposure for occupants within the ICS at 4 ACH compared to the airplane at 100% flow rate.

Outliers in the ICS occurred when occupants were exposed to high concentrations of the cough plume prior to dilution with the surrounding air. Direct exposure to such concentrations were not observed in the aircraft simulations due to the forward facing position of passengers (rather than facing each other) and because the high back passenger seats acted as barriers, similar to Plexiglas™ commonly seen in other environments.

The ECS design of an environment has a large impact on the exposure of individual occupants. Aircraft ECS are deliberately designed to limit fore to aft airflow and to rapidly remove air pollutants such as cigarette smoke, or unpleasant odors, with each row containing multiple air inlets and return air grilles. Whereas design constraints for ICS, such as their size and the complexity of their operation, make control and optimization for an individual occupant more difficult. For example, Figure 6 depicts the nonvolatile mass inhaled for each seat in relation to the index cougher for all ICS scenarios at 4 ACH, and three airplane scenarios at 100% flow rate. Each plot in Figure 6 is a different simulation, where the zero position is the index cougher location, the cough is emitted in the direction of the arrow at the index location, each bubble is the position of a single occupant, the bubble size represents the mass inhaled for each occupant, and, for the ICS, the location of the air inlet is indicated.

**Figure 6.**
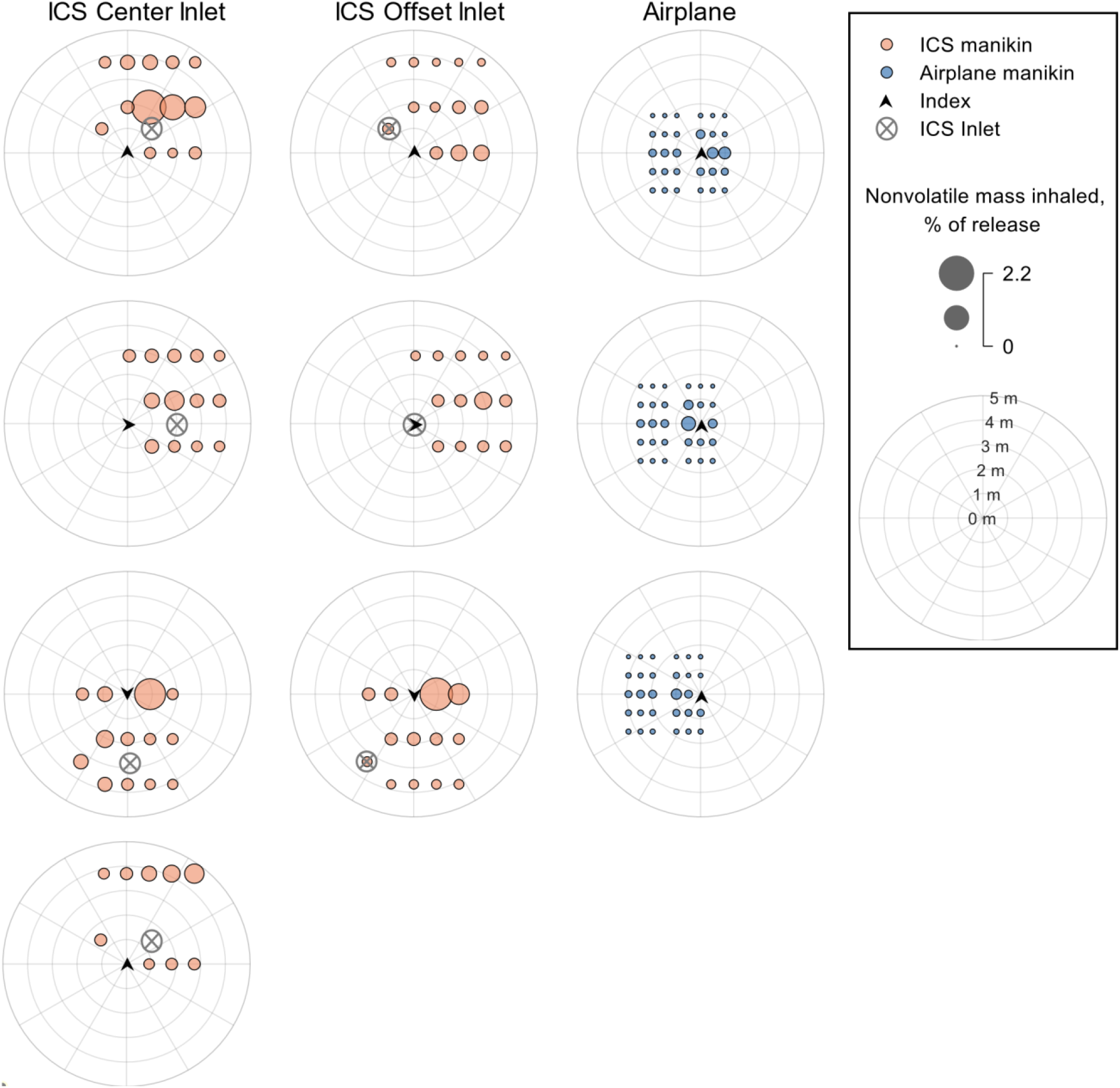
Exposure within the ICS at 4 ACH was higher and more variable than within the airplane at 100% ACH. Polar bubble seat charts showing the mass inhaled as a percent of the mass released for each susceptible occupant in relation to the index location (center of each plot). All ICS simulations at 4 ACH, and three airplane simulations at 100% flow rate are shown. Supply air inlets for the airplane are located above every triplet of seats.

Air inlets on the airplane are located above each triplet of seats (e.g. seats 3A, 3B, and 3C), and are not shown in Figure 6. In the ICS, with an index in seat 4, an occupant in seat 7 was exposed to 2.2% of the released mass when the inlet was in the center position, but 0.1% of the released mass with an offset inlet position. Depending on the inlet position and index position, the maximum mass inhaled in the ICS at 4 ACH ranged from 0.5% to 2.2% of the nonvolatile mass released. The direction of the air flow at the location of the index cougher, which varies throughout the ICS, contributes to the direction the cough plume travels resulting in a wide range of exposure levels. Conversely, in an aircraft, the highest exposure during the simulation was by a susceptible passenger within the same triplet as the index for all scenarios, and the maximum mass inhaled ranged from 0.15% to 0.3%

In both the aircraft and ICS environments, the mass inhaled generally decreased as the distance increased between the index cougher and susceptible inhaler (Supplementary Figure S2). However, exposure to the cough plume in the ICS, and the row by row seating with limited fore to aft flow in the airplane complicated this trend. For an index subject in seat 4, a susceptible occupant 2.1 meters away from the index, seat 7, was exposed to 2.2% of the cough. However, an occupant 0.9 meters away, seat 3, was exposed to 0.18% of the cough. The variability in mass inhaled versus distance in the ICS was due to entrainment of the cough plume in the surrounding air flow, directing the plume towards different locations. For the airplane, the limited fore/aft air flow resulted in variability in mass inhaled with distance. For example, with an index in seat 3F, the mass inhaled at seat 3D, 1 meter away from the index, was 0.15% of the mass released. However, at 0.9 meters away from the index seat 2E was exposed to 0.015% of the mass released. Despite the closer distance to the index, the occupant in seat 2E was exposed to less of the cough than seat 3D because seat 3D is within the same triplet as the index, and seat 2E is in a different row. Despite the close-quarters seating on an aircraft, exposure decreases rapidly as distance increases from the index cougher, particuarly for manikin positions not within the same triplet as the index.

### Limitations

The cases studied in the present work represented typical aircraft cabin and ICS environments. However, other factors that are expected to affect particulate transmission have not been studied, either here or elsewhere. The limitations for the aircraft cabin model are well covered in our earlier publication [2] and thus are omitted here. The following are limitations of the ICS model. They represent areas where the analysis is limited by the large number of variables that would need to be explored for a complete treatment:

- The ICS configuration affects the airflow pattern which varies throughout the indoor model environment. The ICS can also include configurations that are not part of the original architectural plan and are instead defined by the building operator and its occupants. These configurations include furniture arrangements, changes in occupancy and other temporary changes; all of which may affect the particle content in the breathing zones of susceptible subjects. In an aircraft, the positions of seats are unchanging.
- In the ICS, the temperature and humidity are expected to affect particle dynamics and may also affect the airflow pattern. When the flux of occupants in and out of the indoor space is large, temperature and humidity may deviate significantly from the values studied and do so rapidly. For an aircraft in flight, these changes are slow and less likely to cause large changes in air flow patterns.
- Further, during times when the fluctuation in the number of occupants is high, the doors of the indoor space may be opened repeatedly over a short period of time which may significantly alter the airflow pattern. In addition to airflow, subject movements change the spatial relationships between index and susceptible subjects.
- The use of masks and other face coverings capture a portion of the expiratory material and redirect the flow of the remainder; the ratio between these modes depending largely on the fit of the mask to the subjects face.
- The size of the ICS environment can impact the total mass inhaled by providing more volume for dilution. It is expected that by increasing the volume of the ICS, the minimum and average mass inhaled will decrease. The maximum mass inhaled is not expected to significantly decrease with an increased ICS volume since the maxima occurred when there was direct exposure to the cough plume prior to any significant dilution.

A myriad of factors defined by human behavior may all have an effect on air flow and the plume of a cough by an infectious index cougher. This study was configured as stated in the methods section and the values calculated are for those stated configurations. As a perturbation study, the work presented herein only addresses the exposure over the time period necessary to remove the cough particles from the environment. Using a method to estimate the total mass of inhalation over time for both continuous (e.g., breathing or talking) and discontinuous (e.g., coughing) processes may improve our understanding of the risk of infection. However as of yet, the number of viruses per mass exhaled, that is required to cause an infection, has not been determined for COVID-19 with sufficient rigor. Estimates of infectious dose still vary by at least two orders of magnitude and may be defined more by the variation in human biology than by other factors.

### Recommendations for Future Work

In the future, the study of the many factors that may influence the spread of respiratory disease in both indoor commercial and aircraft environments should be further pursued. Those studies might include the analysis of continuous processes like breathing and talking, both of which were considered while preparing the study presented herein. However, the mass of particles released over time during breathing and talking, were deemed smaller than that for a cough thus the focus of the data presented herein.

## Conclusion

The high level of control present in an aircraft is not present in most indoor spaces as demonstrated by the relatively large variation in CFD data for the ICS presented herein. In ICS environments, the people that populate them may or may not be facing one another and they may or may not be downwind from another individual. Overall, as demonstrated in this study, persons in the ICS will experience a higher concentration of particles over a longer period of time due to lower air exchange rates, and a less well defined and controlled direction of air movement than can be expected in an aircraft. Conversely, people within an ICS have more freedom to exit the facility at any time, which is not possible in an aircraft.

This paper compared the exposure from a single cough in an aircraft cabin to the same cough in an ICS. To do this, the mass of particles inhaled was estimated using CFD for both environments and a ratio was calculated comparing the average, minimum, and maximum mass inhaled. The maximum mass of expiratory particles that a passenger on an aircraft inhales after a cough by a nearby passenger is approximately seven times less than the maximum inhaled by a person in a typical ICS. Additionally, when compared to the ICS simulated at 4 ACH, the aircraft achieved 80% particle removal 5 to 12 times more rapidly.

The rapid removal of particles and reduced probability of inhaling contaminated particles is due to the engineering controls on modern aircraft that include: airflow higher than typical ICS, directional downward-flow ventilation designed to minimize fore to aft airflow, HEPA filtration that removes 99.97% of particles from supplied air, separation of rows by high back seats, and the positioning of passengers so that they do not face one another for the majority of the flight. The ventilation system in the aircraft provides an engineered physical distance for adjacent seats, adding an important layer to the overall approach adopted by the society at large to protect against airborne transmission of disease. The effects of other layers in the system of protections (e.g., masks) are treated herein as independent and should be applied in unison with the protection afforded by aircraft ECS system design and performance.

## Supporting information

Supplemental Data for Figures

## Data Availability

All inputs and outputs reported in this paper are provided either in the main text or as
Supplementary Information, which includes an Excel file of tabular data. Researchers
are encouraged to contact the corresponding author for clarifications, advice on data
utilization, and collaborations. All data used to generate plots has also been archived at FigShare.

https://figshare.com/articles/dataset/Comparison_of_cough_particle_exposure_for_indoor_commercial_and_aircraft_cabin_spaces/14292941

## Data Availability

All inputs and outputs reported in this paper are provided either in the main text or as Supplementary Information, which includes an Excel file of tabular data. Researchers are encouraged to contact the corresponding author for clarifications, advice on data utilization, and collaborations.

## Acknowledgements

The authors wish to thank Ted Wu, Matt Schwab, Chao-Hsin Lin, Malia Zee, Steve Baughcum, and Robert Atmur for many productive discussions. The authors thank the High Performance Computing staff at Boeing who prioritized access to computing resources for this activity and provided around-the-clock server support. The authors acknowledge Ansys for providing licenses for their Fluent software to support Confident Travel Initiative studies at Boeing. Finally the authors thank the Boeing technical writing staff for all their help in making this paper more readable and understandable.

## Author Contributions

ACD, ADC, DJM and NAO designed the experiments and set the methodologies.

ADC, ACD and NAO provided parameterization prior to computational fluid dynamics calculations being made.

DJM ran all computational fluid dynamic models and made modifications as needed for each case studied.

ADC created the layout of the indoor commercial space and calculated the heat load balance to define all boundary conditions.

ACD, DJM and NAO performed data analysis and validated the data content. ACD, and NAO wrote sections of the original draft and edited the document.

ACD, ADC, DJM and NAO curated the data.

ACD and NAO wrote and edited the original draft and all subsequent versions of the paper.

ADC and JJC provided resources including access to off-the-shelf software, software licenses, configuration parameters, labor, and computational resources.

JJC provided supervision, administration, and acquisition of funding.

NAO as corresponding author ensured that original data upon which the submission is based is preserved and retrievable for reanalysis, approved data presentation as representative of the original data, and has foreseen and minimized obstacles to the sharing of data described in the work.

## Competing Interests

All authors are employees of The Boeing Company and have no other competing interests. Funding and other resources for this work were provided by The Boeing Company. In-kind contribution was provided by Ansys through supply of licenses for Fluent software.

## Notes

### Author Declarations

No human subjects were used in this study.

