## Supplemental Figures and Tables for "Comparison of cough particle exposure for indoor commercial and aircraft cabin spaces"

#### File Contents

Figure S1. Breathing zone definition in front of the face of a manikin. ........................................S2

Figure S2. Exposure on an airplane is reduced more rapidly with distance than in an ICS. Nonvolatile mass inhaled (% of release) versus distance from the index seat for all airplane simulations at 100% flow rate, and all ICS simulations at 4 ACH on a semi-log scale ...............S2

Table S1. Case summary for aircraft cabin simulations see reference (2) for complete description.........................................................................................................................S3

Table S2. Case summary for ICS simulations. ..............................................................................S4

#### Other Supplementary Materials

Tabular data for Figure 4.

Tabular data for Figures 5, 6 and S2.

### Supplementary Figures

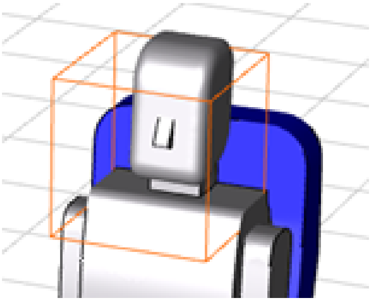

Figure S1. Breathing zone definition in front of the face of a manikin.

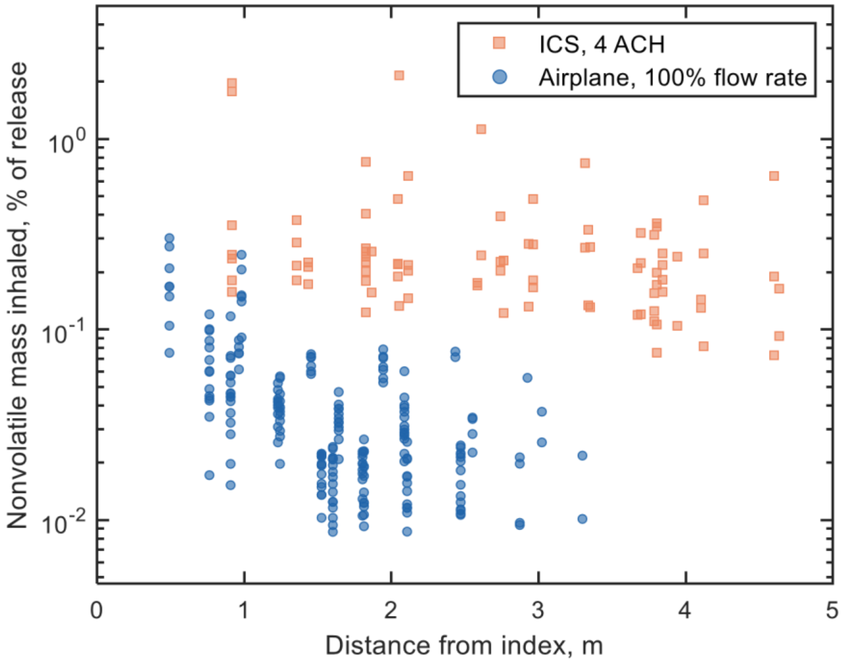


Figure S2. Exposure on an airplane is reduced more rapidly with distance than in an ICS. Nonvolatile mass inhaled (% of release) versus distance from the index seat for all airplane simulations at 100% flow rate, and all ICS simulations at 4 ACH on a semi-log scale.

#### Supplementary Tables

Table S1. Case summary for aircraft cabin simulations see reference (2) for complete description.

| Index Cougher Seat | Initial Condition | Flow Rate | Flow Rate (ACFM) | ACFM per Occupant | Relative Humidity |
|---|---|---|---|---|---|
| 3D | No offset | 100% | 588 | 20 | 0% |
| 3D | 90 seconds offset | 100% | 588 | 20 | 0% |
| 3D | 120 seconds offset | 100% | 588 | 20 | 0% |
| 3E | No offset | 100% | 588 | 20 | 0% |
| 3F | No offset | 100% | 588 | 20 | 0% |
| 3D | No offset | 77% | 453 | 15 | 0% |
| 3D | No offset | 55% | 323 | 11 | 0% |
| 3D | No offset | 100% | 588 | 20 | 10% |
| 3D | No offset | 100% | 588 | 20 | 20% |

Table S2. Case summary for ICS simulations.

| Index Cougher Seat | Inlet Position | Flow Rate (ACFM) | Flow Rate (ACH) | ACFM per Occupant |
|---|---|---|---|---|
| 4 | Center | 683 | 8 | 49 |
| 4 | Center | 342 | 4 | 24 |
| 4 | Center, Removed Seats 6-9 | 342 | 4 | 34 |
| 4 | Center | 171 | 2 | 12 |
| 4 | Offset | 342 | 4 | 24 |
| 5 | Center | 342 | 4 | 24 |
| 5 | Offset | 342 | 4 | 24 |
| 12 | Center | 342 | 4 | 24 |
| 12 | Offset | 342 | 4 | 24 |
